# Interviewer Biases in Medical Survey Data: The Example of Blood Pressure Measurements

**DOI:** 10.1101/2023.04.11.23288399

**Authors:** Pascal Geldsetzer, Andrew Young Chang, Erik Meijer, Nikkil Sudharsanan, Vivek Charu, Peter Kramlinger, Richard Haarburger

**Affiliations:** Division of Primary Care and Population Health, Department of Medicine, Stanford University, Stanford, CA, USA; Department of Epidemiology and Population Health, Stanford University, Stanford, CA, USA; Chan Zuckerberg Biohub, San Francisco, CA, USA; Stanford Cardiovascular Institute, Stanford University, Stanford, CA, USA; Center for Innovation in Global Health, Stanford University, Stanford, CA, USA; Center for Economic and Social Research, University of Southern California, Los Angeles, CA, USA; Assistant Professorship of Behavioral Science for Disease Prevention and Health Care, Technical University of Munich, Munich, Germany; Quantitative Sciences Unit, Department of Medicine, Stanford University, Stanford, CA, USA; Department of Pathology, Stanford University, Stanford, CA, USA; Department of Statistics, University of California Davis, Davis, CA, USA; Research Training Group “Globalization and Development”, Goettingen University, Goettingen, Germany

**Keywords:** Blood Pressure, Hypertension, Measurement Error, Public Health, Health Survey

## Abstract

Health agencies rely upon survey-based physical measures to estimate the prevalence of key global health indicators such as hypertension. Such measures are usually collected by non-healthcare worker personnel and are potentially subject to measurement error due to variations in interviewer technique and setting, termed “interviewer effects”. In the context of physical measurements, particularly in low- and middle-income countries, interviewer-induced biases have not yet been examined. Using blood pressure as a case study, we aimed to determine the relative contribution of interviewer effects on the total variance of blood pressure measurements in three large nationally-representative health surveys from the Global South.

We utilized 169,681 observations between 2008 and 2019 from three health surveys (Indonesia Family Life Survey, National Income Dynamics Study of South Africa, and Longitudinal Aging Study in India). In a linear mixed model, we modeled systolic blood pressure as a continuous dependent variable and interviewer effects as random effects alongside individual factors as covariates. To quantify the interviewer effect-induced uncertainty in hypertension prevalence, we utilized a bootstrap approach comparing sub-samples of observed blood pressure measurements to their adjusted counterparts. Our analysis revealed that the proportion of variation contributed by interviewers to blood pressure measurements was statistically significant but small: approximately 0.24-2.2% depending on the cohort. Thus, hypertension prevalence estimates were not substantially impacted at national scales. However, individual extreme interviewers could account for measurement divergences as high as 12%. Thus, highly biased interviewers could have important impacts on hypertension estimates at the sub-district level.

**Significance Statement:** Physical measurements such as blood pressure are important indicators of countries’ health system performance. These measures are usually obtained in household surveys by study-specific interviewers, who are not clinical healthcare workers. Thus, there is a concern that they may contribute substantial measurement error. We used three large, nationally-representative health surveys from India, Indonesia, and South Africa to model the magnitude of the interviewer effect on blood pressure measurements, then projected their impact on estimations of country-level hypertension prevalence. At smaller geographic units, “extreme” interviewers could substantially bias hypertension estimates. Overall, however, the magnitude of the interviewer effects was small and, thus, unlikely to substantially bias hypertension prevalence estimates at the national level.

## 1 Introduction

Global health indicators such as blood pressure, weight, and height are critical for monitoring both national and international health system performance. Such markers are largely collected through household surveys, which are often seen as the gold standard methodology due to their population-representative nature (Ties Boerma and Sommerfelt, 1993; Corsi et al., 2012; Boerma et al., 2003; Clark and Sanderson, 2009; Mbondji et al., 2014).

Interviewer-collected physical measures such as heart rate or body mass index (BMI) may appear to hold greater “objectivity” than self-reported indicators or subjective social indicators. Self-reported data is frequently prone to not only random measurement error, but also systematic measurement error due to interviewee attitudes such as recall bias and social desirability bias (Althubaiti, 2016). Nevertheless, physical measures are still subject to a substantial degree of random measurement error due to administrator technique and environmental context during acquisition (Ulijaszek and Kerr, 1999; Cernat and Sakshaug, 2020; Ali and Rouse, 2002; Svensson and Theorell, 1982). This phenomenon may possibly be magnified in the case where medical measurements are taken by non-clinician interviewers who may not routinely perform such measures outside of the research setting.

Nevertheless, many household surveys make the implicit assumption that, after their training, interviewers all perform to the same standard as one another (Jaszczak et al., 2009). Subsequent analyses therefore assume that the interviewers are not a source of measurement error and that uncertainty estimates are purely based on the sampling strategy.

At the national level, these “interviewer effects” may average out from the large number of interviewers contributing both positive and negative measurement error. At finer geographic divisions, however, the relatively smaller number of interviewers may lead to greater variation or even potential bias in the measurement of a target indicator. This is particularly important because estimates from small areas are increasingly being used in public health decision making and for mapping disease prevalence at subnational levels, sometimes in resolutions as fine as 5 × 5 km (Dwyer-Lindgren et al., 2019; Reiner Jr et al., 2018; Osgood-Zimmerman et al., 2018; Graetz et al., 2018).

Prior analyses have queried the intra- and inter-observer reproducibility of specific physical measures, but such investigations have tended to focus on the reliability of these markers for clinical situations (Ali and Rouse, 2002; Svensson and Theorell, 1982; Schulze et al., 2000). Furthermore, most such studies have utilized healthcare workers like nurses and medical trainees as the measurement-takers given their applicability to the medical setting, and have examined high-income country populations (Bogan et al., 1993; Dickson and Hajjar, 2007). Large-scale empirical analyses of non-clinician interviewers’ reliability for physical measures for public health purposes, especially in low- and middle-income countries (LMICs), remain sparse. The amount of random measurement error found in such global health indicators varies, with some exhibiting relatively low degrees (e.g., controlled laboratory-based tests) while others with increased operator inputs suffer from potentially greater degrees of interviewer-introduced measurement error. For example, anthropometry for newborns, adult waist circumference, and blood pressure measurements require interviewers to make subjective decisions about how and where to place the instruments and in what settings to do so (Cernat and Sakshaug, 2020, 2021).

Here we assess the magnitude of interviewer-induced measurement error in large-scale global health surveys using the case study of high blood pressure. High blood pressure is an ideal case study because it is already a disease of considerable importance in low- and middle-income countries (LMICs) (Zhou et al., 2017; Yusuf et al., 2020). Blood pressure is readily and frequently measured noninvasively, and non-clinician study personnel can be taught how to collect blood pressure assessments (Jaszczak et al., 2009). This is particularly important as community health workers and other non-nurse/non-physician healthcare workers are increasingly being called upon to care for noncommunicable diseases in primary care in poor countries, and they are also frequently called upon for survey data collection as well (Jeet et al., 2017; Singh and Sachs, 2013; Otieno et al., 2012).

As such, in the present analysis (assuming that interviewers are randomly allocated to house-holds within primary sampling units) we examine the magnitude of uncertainty attributable to interviewer effects on blood pressure measurements and hypertension (systolic blood pressure ≥ 140*mmHg*) in three large longitudinal health surveys from the Global South.

## 2 Results

### 2.1 Sample Characteristics

Table 1 shows descriptive statistics for the data sets used in this study after pre-processing. Data from 169,681 total encounters were utilized, with 26,554 from the Indonesia Family Life Survey (IFLS), 55,469 from the Longitudinal Aging Study in India (LASI), and 87,658 from the National Income Dynamics Study (NIDS) of South Africa, respectively.

**Table 1:** Descriptives of IFLS, NIDS and LASI data.

|  | IFLS<br>(N=26554) | LASI<br>(N=55469) | NIDS<br>(N=87658) | Overall<br>(N=169681) |
| --- | --- | --- | --- | --- |
| **Average SBP measurement** |  |  |  |  |
| Mean (SD) | 129 (19.9) | 129 (19.1) | 121 (21.3) | 125 (20.8) |
| Median [Min, Max] | 126 [68.0, 241] | 127 [60.0, 234] | 117 [44.0, 240] | 122 [44.0, 241] |
| **Sex** |  |  |  |  |
| Male | 16569 (62.4%) | 25694 (46.3%) | 36446 (41.6%) | 78709 (46.4%) |
| Female | 9985 (37.6%) | 29775 (53.7%) | 51212 (58.4%) | 90972 (53.6%) |
| **Age** |  |  |  |  |
| Mean (SD) | 41.7 (13.1) | 59.5 (10.4) | 36.1 (17.1) | 44.6 (18.0) |
| Median [Min, Max] | 42.0 [15.0, 101] | 58.0 [45.0, 108] | 32.0 [14.0, 108] | 46.0 [14.0, 108] |
| **Education** |  |  |  |  |
| Less than primary | 0 (0%) | 6317 (11.4%) | 8224 (9.4%) | 14541 (8.6%) |
| Primary or secondary | 22629 (85.2%) | 42613 (76.8%) | 67901 (77.5%) | 133143 (78.5%) |
| Tertiary | 3925 (14.8%) | 6539 (11.8%) | 11533 (13.2%) | 21997 (13.0%) |
| **Body-mass-index (BMI)** |  |  |  |  |
| Mean (SD) | 23.3 (4.28) | 22.8 (4.73) | 26.1 (6.72) | 24.6 (6.00) |
| Median [Min, Max] | 22.7 [10.7, 57.1] | 22.3 [10.5, 55.6] | 24.6 [10.4, 60.0] | 23.4 [10.4, 60.0] |
| **Ever diagnosed with hypertension** |  |  |  |  |
| Not diagnosed | 23406 (88.1%) | 39600 (71.4%) | 75814 (86.5%) | 138820 (81.8%) |
| Diagnosed | 3148 (11.9%) | 15869 (28.6%) | 11844 (13.5%) | 30861 (18.2%) |
| **Log income** |  |  |  |  |
| Mean (SD) | 12.5 (4.04) | 11.2 (1.66) | 7.95 (0.942) | 9.71 (2.72) |
| Median [Min, Max] | 13.7 [0, 19.5] | 11.4 [0, 18.8] | 7.82 [4.25, 13.0] | 9.13 [0, 19.5] |
| **Smoking** |  |  |  |  |
| Non-smoker | 14850 (55.9%) | 47686 (86.0%) | 75814 (86.5%) | 138350 (81.5%) |
| Smoker | 11704 (44.1%) | 7783 (14.0%) | 11844 (13.5%) | 31331 (18.5%) |
| **Urban/Rural** |  |  |  |  |
| Urban | 15300 (57.6%) | 19263 (34.7%) | 43477 (49.6%) | 78040 (46.0%) |
| Rural | 11254 (42.4%) | 36206 (65.3%) | 44181 (50.4%) | 91641 (54.0%) |

### 2.2 Variation shares in hypertension prevalence

To interpret the effect sizes of the interviewer level-effects, we compare their shares in total variation to the shares of other level-effects and the residual from the same estimations. Table 2 presents the variance components of the fitted linear mixed models (LMM) for the IFLS, NIDS, and LASI datasets.

**Table 2:** Variance components of the fitted LMMs by data set.

|  | Effect | Variance | Percentage |
| --- | --- | --- | --- |
| **IFLS** |  |  |  |
|  | Household | 37.6 | 13.6% |
|  | Interviewer | 1.47 | 0.53% |
|  | Province | 2.96 | 1.07% |
|  | Municipality | 2.29 | 0.83% |
|  | Subdistrict | 2.78 | 1.01% |
|  | Residuals | 229 | 82.9% |
| | Total | 276.1 | $\approx 100\%$ |
| **NIDS** |  |  |  |
|  | Household | 39.6 | 12.1% |
|  | Cluster | 3.74 | 1.15% |
|  | Interviewer | 7.19 | 2.2% |
|  | Province | 3.06 | 0.94% |
|  | Residuals | 273 | 83.7% |
| | Total | 328.17 | $\approx 100\%$ |
| **LASI** |  |  |  |
|  | Household | 21.3 | 6.55% |
|  | Interviewer | 0.785 | 0.24% |
|  | State | 7.99 | 2.46% |
|  | District | 7.75 | 2.39% |
|  | Village/ward | 4.96 | 1.53% |
|  | Residuals | 282 | 86.8% |
| | Total | 324.785 | $\approx 100\%$ |

The bootstrap likelihood ratio test (LRT) tests give p-values of p < 0.0001 for all three datasets. This strongly suggests the presence of interviewer effects in all three datasets, although they are numerically small.

### 2.3 Uncertainty in Sample Hypertension Prevalence

Figure 1 displays the non-parametric bootstrap densities for hypertension prevalence, based on the original data (blue, dashed), and the corrected measurements (red, dotted). The vertical line represents the observed prevalence by data source.

**Figure 1:**
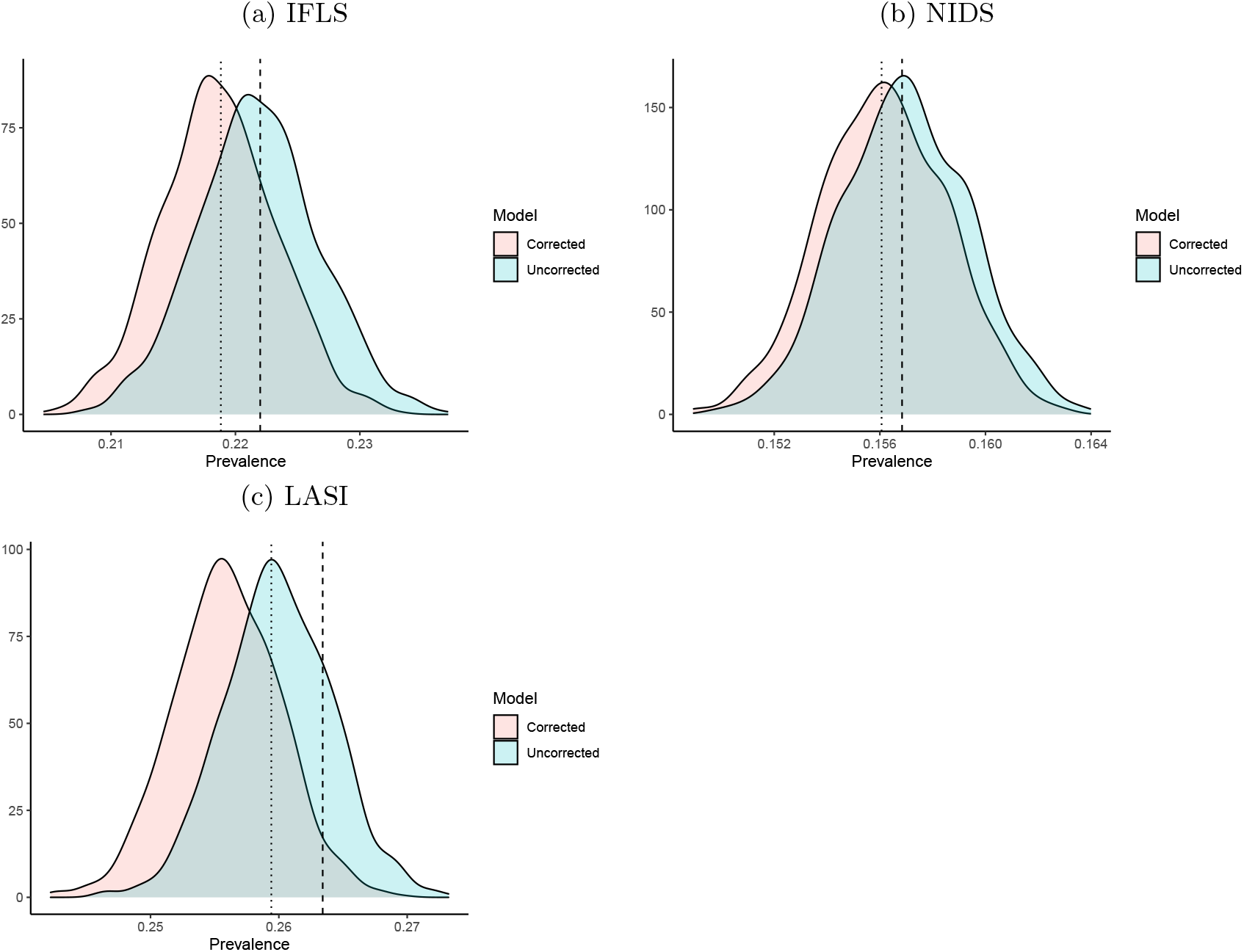
Bootstrap densities for hypertension prevalence, based on the original data (blue, dashed), and the corrected measurements (red, dotted). The vertical line represents the observed prevalence.

### 2.4 Effect Study

In order to illustrate the interviewer-introduced uncertainty in hypertension prevalences, we perform an effect study. Using the set of observed systolic blood pressure measurements and the measurements corrected for the estimated interviewer effects, we can compare observed interviewer-specific prevalences of hypertension to the respective corrected interviewer-specific prevalences. Alternatively, we can also illustrate differences in prevalences for geographic areas, such as sub-districts.

#### 2.4.1 Interviewer-Specific Prevalences: Observed and Corrected

Figure 2 illustrates a sub-sample of the interviewer-specific observed and adjusted prevalences of hypertension for the IFLS dataset. The sub-sample is created based on the distribution of differences in observed and adjusted prevalences. For example, to focus on the most extreme cases, we depict the prevalences for all interviewers for whom the difference between observed and adjusted prevalence lies above the 70th-percentile of these differences. In other words, we show the 30 percent of cases subject to the most drastic adjustment effects. The top 50%, 30%, 10%, and 1% cases are presented.

**Figure 2:**
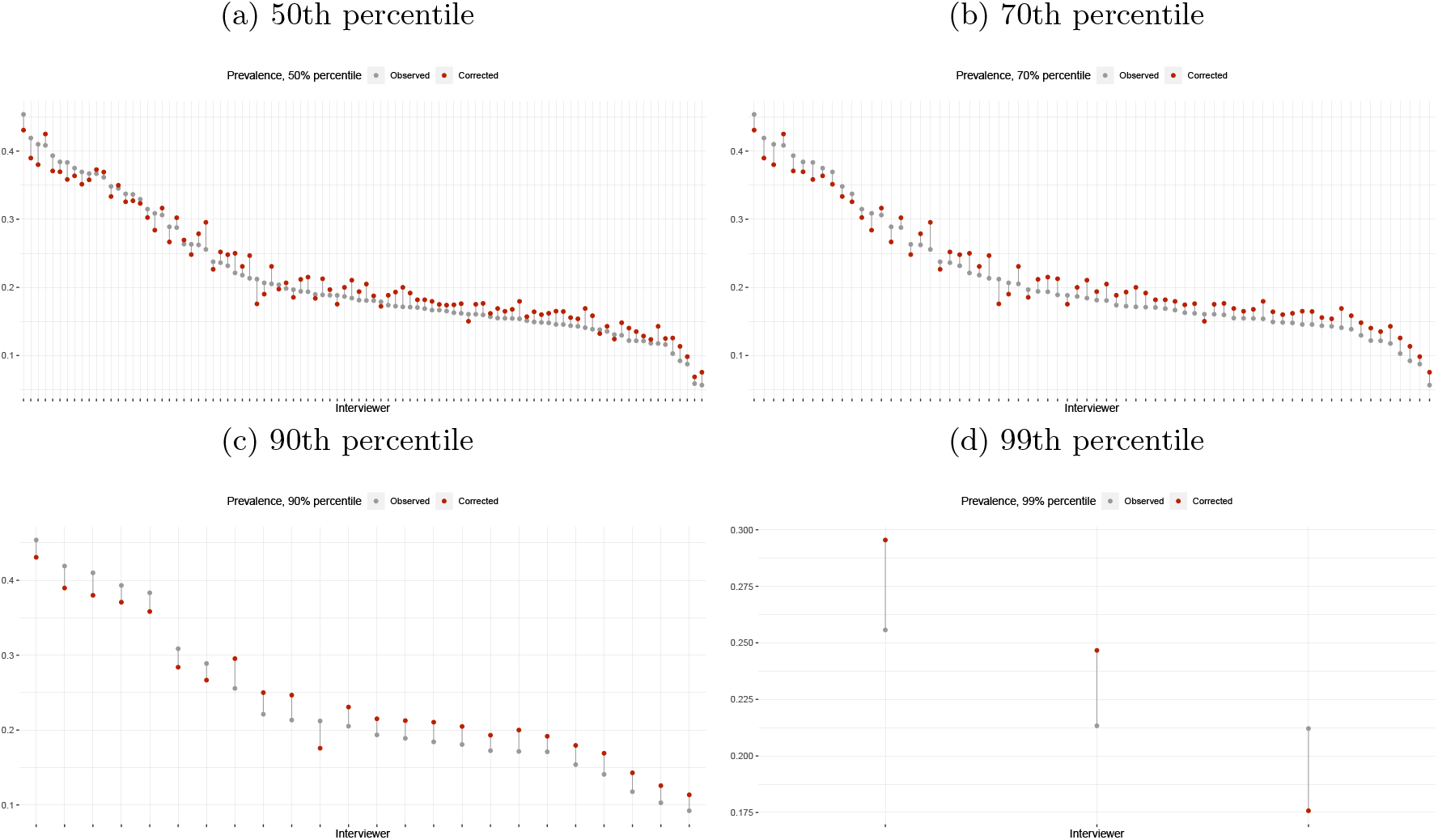
IFLS: Observed and adjusted interviewer specific prevalences of hypertension, 50%, 30%, 10%, 1% of cases subject to largest adjustment effects.

The analogous findings for NIDS and LASI are presented as Supplementary Material 1.

#### 2.4.2 Sub-district specific prevalances: observed and corrected

Analogously to the interviewer specific prevalances, we can also depict changes in prevalances for geographical units, as illustrated in Figure 3. The higher the granularity in geographical division, the larger the influence of single interviewers. We thus depict adjustment induced changes in prevalences on the most granular level available for each respective data set. In case of LASI and IFLS the most granular geographical level are sub-districts. In the case of NIDS less granular level data is available, so that we are limited to the cluster level.

**Figure 3:**
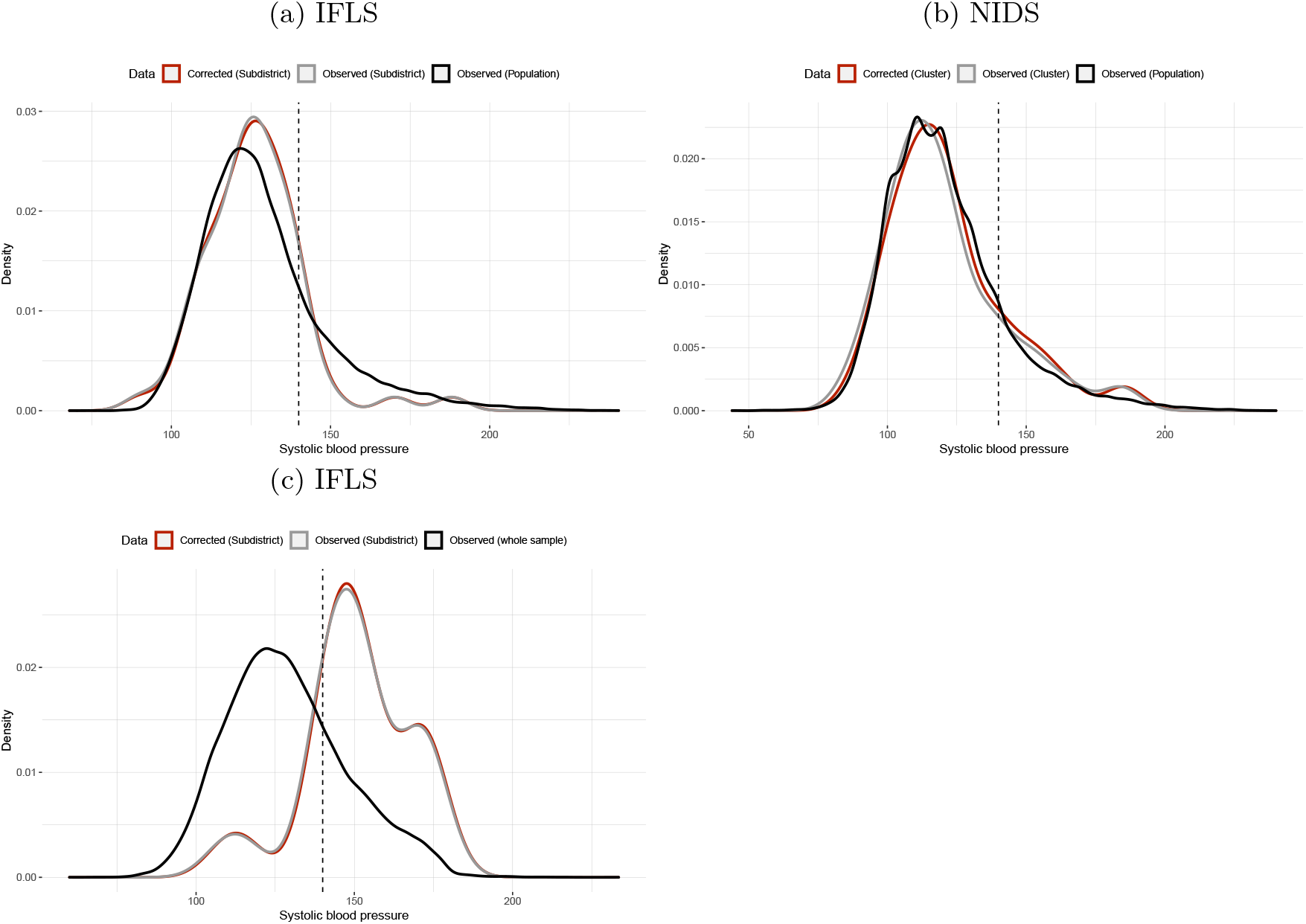
Systolic blood pressure densities, observed and adjusted for estimated interviewer effects, for selected subdistricts subject to large adjustment induced changes by data source. Population densities are added as comparison.

## 3 Discussion

In the present analysis, we found that interviewer effects in blood pressure measurements were statistically significant, although numerically trivial, in three large longitudinal health surveys from Indonesia, India, and South Africa. This was achieved by calculating the proportion of total variance attributable to various sources, one of which was the interviewer. Nevertheless, both the absolute and relative contribution of the interviewer to blood pressure measurement variation was not particularly high, especially when compared to geographic/community-level effects. In the IFLS cohort, interviewer-level effects comprised 0.5% of the variance, while in NIDS, 2.2%, and in LASI, 0.2%. In fact, household effects (13.6%, 12.1%, 6.6%, respectively) dominated the variance of all three datasets, with residential effects (i.e., province, state, subdistrict, municipality) higher than interviewer effects except for in NIDS.

On the population level, however, the combined interviewer effect could potentially impact the uncertainty in hypertension prevalence. As such, we generated non-parametric bootstraps of prevalence estimates unadjusted and adjusted for the interviewer effect, which show very small but consistently lower point estimates of hypertension prevalence in all three datasets on the order of a fraction of a percent. This may have minor implications for public policy targeting hypertension and suggest slight present overestimation of true hypertension prevalence in these settings.

Nevertheless, the magnitude of the discrepancies is not exceedingly high at these larger scales — where we found the interviewer effect to carry the greatest possibility of influencing hypertension estimation was at smaller geographic divisions. Taking the most “extreme” individual interviewers responsible for the greatest adjustment effects in each dataset and comparing their observed and adjusted hypertension prevalences revealed divergences as high as 12% in NIDS. We therefore assessed their impacts by comparing the observed and interviewer-effect adjusted sub-district specific hypertension prevalences subject to the greatest adjustment effects. These revealed up to 5-7 percentage points (p.p.) prevalence differences between observed and corrected values at sub-district levels for the top 1% of cases subject to adjustment effects. The substantial degree of bias that these may introduce at the local level compared to the population (or whole sample) level are well-visualized in the resultant cluster-specific blood pressure density plots. For example, in LASI, the modal systolic blood pressure signed difference between subdistrict and total population was nearly 25*mmHg*.

Our study represents the largest empirical estimation of interviewer effects on blood pressure. We also believe it to be the first of its kind involving low- and middle-income country populations. Thus, it contributes to the growing body of work examining and quantifying interviewer-based sources of measurement error for survey-based global public health indicators. The results are reassuring that the present strategy of utilizing non-clinician study interviewers is likely not generating a critical degree of variation in blood pressure measurement for populations, and we propose one possible method by which analysts may adjust for these small interviewer effects.

Because our investigation is, to our knowledge, the first to assess interviewer-effects for blood pressure in household surveys from low- and middle-income countries, we are only able to compare our findings to those from much smaller samples in two surveys from the UK. Cernat and Sakshaug found that there are interviewer effects on measurement error from both nurses and trained non-clinician interviewers in these two UK-based surveys (Cernat and Sakshaug, 2020, 2021). For non-clinician interviewers, they noted that in measures such as height, weight, blood pressure, and pulse, interviewer effects similarly comprised only a small fraction of the variance—for blood pressure, less than 1%. Much like our findings, these studies also identified that area-level effects contributed a greater source of variation than the interviewer effect for many physical measures.

Nevertheless, our work further models the public health implications of the interviewer effect by estimating the impact of these forces on hypertension prevalence estimates at multiple geographic levels. In doing so, our analyses also identified that extremely biased interviewers could lead to markedly biased hypertension estimates, and that if there is disproportionate allocation of these “extreme” interviewers to a locale at the level of a sub-district or smaller, that there may be substantially biased hypertension prevalence estimates in these geographic units.

Strengths of our study include the size of the analytic cohort (total 169,681 observations), as well as the use of three different nationally-representative datasets from Africa, South Asia, and Southeast Asia. There is substantial heterogeneity in the resultant populations, not just by the distribution of gender, age, and urban/rural breakdown, but also the underlying true prevalence of hypertension. Blood pressure measurements from years 2008 through 2019 were included, further capturing time-related variation. The most important limitation of our analysis is that, analytically, our modeling strategy relies upon the assumption that all interviewers were quasi-randomly allocated to participants within the primary sampling units. Other limitations of the work include the inclusion of systolic blood pressure only, for reasons of statistical feasibility. Diastolic hypertension (both independently and in conjunction with systolic hypertension) may be a risk factor for adverse cardiovascular outcomes (Flint et al., 2019; Strandberg et al., 2002). In addition, the LASI cohort was substantially older than the IFLS and NIDS cohorts. Furthermore, the full dataset does not constitute a random sample of all household surveys in low- and middle-income countries. Lastly, all three survey cohorts involved interviewers who were highly trained using established, high-quality protocols and closely monitored by study administration. As biased interviewers have higher impact on measurement error in small geographic units, our results may underestimate the magnitude of interviewer effects for less-rigorously trained/observed interviewers in LMIC settings.

We conclude by noting that interviewer effects appear to be present, but small at best in household surveys of blood pressure in lower middle- and middle-income countries. Future work could involve targeted empirical analyses of the influence of “extreme interviewers” on quantifying the local burden of disease, as well as replication of our methods in other cohorts from different continents and from low-income countries. Additionally, we recognize that blood pressure is but one physical measure from a large pool of monitored global health indicators. As prior research in other settings has suggested that interviewer effects vary with the type of measurement performed, independent analyses of these other markers such as weight and body mass index should be pursued to provide a more comprehensive understanding of the phenomenon.

## 4 Materials and Methods

### 4.1 Data Sources

We demonstrate the implications of interviewer measurement biases using three common longitudinal health surveys. Besides waves 4 and 5, as well the east extension of the Indonesia Family Life Survey (IFLS), we use all five waves of the National Income Dynamics Study (NIDS), and the first wave of the Longitudinal Aging Study in India (LASI) in our analysis. All three data sets were collected with the purpose to document socioeconomic and health outcomes over time. Moreover, they were designed to provide sufficient sample size and adequate sampling schemes to be nationally representative. Thus, they are generally considered suitable to estimate prevalences of diseases for whose documentation adequate examinations were conducted as part of the survey, such as hypertension.

### 4.2 Sampling strategy

NIDS: The NIDS data were collected in five waves between February 2008 and December 2017 (Southern Africa Labour and Development Research Unit, 2018a,b,c,d,e). Since the NIDS data are of longitudinal nature, the households interviewed in the first wave were re-contacted for the following waves. However, the sample was topped up throughout the following waves to account for under-sampled socioeconomic groups and attrition. A two-stage stratified cluster sample design was applied in the data generation process of the first wave.

The underlying 2003 master data used to generate NIDS were provided by Statistics South Africa, comprised 3000 primary sampling units (PSUs), and were stratified with respect to 53 district councils. The NIDS data depict a subset of 400 PSUs which were randomly drawn within the strata, whilst conserving proportionality. Within each PSU, 8 non-overlapping samples of dwelling units had been drawn for the creation of the master data, which are referred to as clusters in the NIDS documentation. The majority of clusters were assigned various household surveys before the creation of NIDS. Two clusters in each PSU however had never been involved in surveys, and became the base for NIDS. For further details see (Leibbrandt et al., 2009). NIDS wave 1 comprises completed surveys of 7,296 households from the aforementioned sub-sampled 400 PSUs. In order to establish national representativeness, different sets of weights were constructed as described in (Wittenberg, 2009). Since our analysis does not aim for national representativeness, but focuses on interviewer effects only, we do not apply the weights provided within the NIDS data and thus do not further discuss the computation of the weights here. Thus, they do not account for the interviewer effects, but merely the sampling weights.

After cleaning and preprocessing the NIDS data as outlined above, 87,658 observations remain, which we use throughout our analysis.

IFLS: The IFLS data used in the scope of this analysis comprise waves four, five and the east extension (Strauss et al., 2009; Sikoki et al., 2013; Strauss et al., 2016). As is the case with NIDS, due to the IFLS data being a longitudinal survey, the households interviewed during the first wave were re-contacted for all following waves. Thus, the sampling scheme of the first wave determined the sample composition of all following waves. IFLS1 stratified on provinces and urban versus rural locations within which simple random sampling was applied. Out of a total of 27 Indonesian provinces, only 13 are included in the sample, which however represented 83% of the population in 1993 (Strauss et al., 2016). Within the selected 13 provinces, 321 enumeration areas (EAs) were randomly chosen, with proportions being selected to cause oversampling of urban EAs and smaller provinces to ensure the comparability of rural and urban EAs. While within each urban EA 20 households were selected, 30 were selected within each rural EA, resulting in a total of 7,224 completed household interviews in IFLS1. For a more detailed description of the sampling scheme please refer to (Strauss et al., 2016). IFLS East includes most of the provinces not covered by the main IFLS. Within each selected province, 14 villages or urban villages were randomly drawn. These were then subdivided into units/areas with about 100-150 households, from which one was drawn at random. Within each of these, again 20 households were drawn if urban and 30 if rural. See (Sikoki et al., 2013) for more details. After initial data cleaning and processing 26,554 individual level observations from IFLS 4, 5, and East remain, which we use in the scope of this analysis.

LASI: We use the first wave of LASI data which was collected between 2017 and 2019 (International Institute for Population Sciences (IIPS), MoHFW, Harvard T. H. Chan School of Public Health (HSPH) and the University of Southern California (USC), 2020). The sampling scheme applied throughout the LASI data collection followed the 2011 census, and implemented a multistage, stratified cluster sample design. While in the case of urban areas three sampling stages were conducted, four stages were conducted in the case of rural areas. The first stage consisted in the selection of PSUs within states. In the second stage villages were selected in the rural PSUs and wards within the urban PSUs. Stage three included the selection of households in rural areas and the selection of Census Enumeration Blocks (CEBs) in wards. The final and fourth stage applied in urban areas comprised the selection of households. The LASI data used in the scope of this analysis comprise 55,469 observations post pre-processing and cleaning.

### 4.3 Interviewer Training, Characteristics, and Monitoring

NIDS: Interviewer training was held at the same time as the pre-test was conducted, specifics on the training of blood pressure measurements are not documented. The NIDS documentation does not mention specially trained health professionals taking the health measurements as is common in similar surveys. Thus, health measurements have been taken by the interviewer conducting the rest of the household surveys.

With wave five a set of interviewer demographics and experience variables were added to the available data.

The use of paradata was implemented to oversee interviewers and thereby reduce interviewer effects. Precisely, paradata are used to monitor questionnaire duration, refusal rates, magnitude of anthropometric measurement differences between current waves and previous waves, flag extreme BMI measures, and run other similar checks. The checks were taken periodically from about 6 weeks into fieldwork or when there were enough data to estimate meaningful averages. When interviewers’ performance measures were conspicuous they were investigated, retrained, moved to different teams for closer supervision or removed. In some cases the respective households were re-interviewed. The Southern Africa Labour and Development Research Unit (SALDRU) carried out a range of pattern searches and consistency checks on the data during fieldwork to identify interviewer effects and potential general cases of mis-capture.

The NIDS sample used in our analysis comprises a total of 513 distinct interviewers taking blood pressure measurements.

IFLS: Supervisory training was held for all senior personnel. In the case of IFLS5 this training of trainers included reviewing all parts of the survey: household, community-facility, health, computer-assisted personal interview system (CAPI) tracking, and the management information systems used in the scope of the data collection. Household interviewer training was conducted in two phases. Training sessions were divided into two parts, classroom training and field practice. Household interviewers received 19 days of classroom training and 4 days of field practice. The collection of health data was conducted by regular interviewers, i.e., no health professionals were involved in the data collection on site during the interviews. Training for health-related measurements was part of the regular interviewer training. In the case of IFLS4 and IFLS East, the CAPI system had not been implemented yet and blood pressure measurements were conducted by nurses, i.e. professional health workers, and non-professional interviewers, respectively.

The combined IFLS data contains a total of 409 distinct interviewers taking blood pressure measurements.

LASI: A series of manuals were designed to standardize different aspects of surveys conducted in the scope of the LASI data collection. These manuals were instrumental in the training of interviewers. One of the manuals specifically focuses on the physical measures section of LASI and thus includes instructions for the measurement of blood pressure. The training duration of interviewers and health investigators was 35 days, of which five took place in the field. Even though the interviewers were employed via sub-contractors, they were trained by trainers, who themselves were trained by the International Institute for Population Sciences (IIPS). After training was completed, investigators were individually assessed to assure that their work met the requirements previously defined by the manuals.

The LASI sample used in our analysis comprises a total of 504 distinct interviewers taking blood pressure measurements.

### 4.4 Definition of Hypertension, Blood Pressure Measurement

Multiple systolic blood pressure measurements were taken in the scope of all surveys included in this study. In the case of the IFLS and LASI data, three measurements were taken per individual, in the case of the NIDS data only two. In order to mitigate the white coat effect and to average out idiosyncratic fluctuations in measurements, we average the second and third measurement, while disregarding the first in the case of IFLS and LASI. In the case of NIDS, we only consider the second measurement, disregarding the first. Following this procedure, we obtain a single systolic blood pressure value for each interviewee. We consider interviewees to be hypertensive if their resulting single systolic blood pressure measurement is equal to or greater than 140 mm Hg.

Measurements were conducted using an Omron HEM 7121 BP monitor in the case of LASI and an Omron HEM 7203 in the case of IFLS. Information on the exact device used for blood pressure measurement throughout NIDS data collection is not part of the publicly available documentation.

### 4.5 Definition of Covariates

We add covariates to the model, which we consider potential determinants of blood pressure. To keep the results comparable, we use mostly the same set of covariates across all data sets. Besides using interviewees’ sex, age, BMI, and smoking status, we proxy interviewees’ socioeconomic background with income and education. The variables we choose in the respective data sets to compose our income proxy refer to monthly salaries and wages or monthly profits from entrepreneurship for NIDS and IFLS, and the logarithm of total household income for LASI. While the resulting income variables are hardly comparable across data sets, we assume comparability within data sets. To align the information on the education of individuals, we re-code education into three categories, namely less than primary schooling, primary and/or secondary schooling and tertiary education, except in LASI, where we added a fourth category for no schooling. In order to proxy for the possible use of blood pressure lowering medication we include a variable which depicts whether an interviewee has ever been diagnosed with hypertension before.

### 4.6 Statistical Analysis

We model a linear relationship of systolic blood pressure and available covariates. Formally, systolic blood pressure for individual *i* in household *j* at location *k* measured by interviewer *l* is denoted as *Y*_*ijkl*_, so that

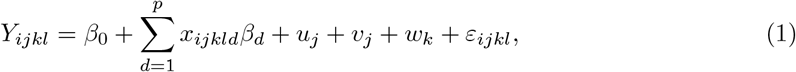

where 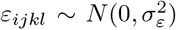 is a Gaussian error term. Furthermore, *x*_*ijkl*1_, …, *x*_*ijklp*_ are the available co-variates, *β*_0_ ∈ ℝ a common intercept and *u*_*j*_, *v*_*k*_, *w*_*l*_ the respective level-effects of household, location and interviewer, for *j* = 1, …, *J, k* = 1, …, *K, l* = 1, …, *L*. These level-effects, as well as the parameter vector *β* = (*β*_0_, *β*_1_, …, *β*_*p*_)^*t*^ are unknown and have to be estimated given a sample of independent measurements.

Since the main objective lies in investigating systolic blood pressure, this model includes a selection of socio-economic control covariates. The separately modeled level-effects include a household effect, the interviewer effect, and the maximum number of geographical level-effects supported by the respective data set. In the following we will motivate the use of the individual level-effects. We suspect that the interviewer effect significantly influences systolic blood pressure measurements, and is at the core of our analysis, as described above. Of note, due to the inability to trace interviewers across waves of the datasets, we treat all observations individually and ignore the time dimension.

We motivate the use of geographical level-effects based on the assumption that geographical cultural clusters, geographical differences in the availability of food, geographical differences in health care access, and similar factors might affect systolic blood pressure spatially.

It is common practice to assign interviewers to households and not to interviewees directly. An interviewer then interviews all eligible individuals belonging to an assigned household. Variation in systolic blood pressure on the household level therefore potentially confounds the estimation of the interviewer effect. Thus, we include household effects to absorb household level variation.

We are interested in investigating *Y*_*ijkl*_ − *w*_*l*_, which is the systolic blood pressure adjusted for the true measurement error induced by interviewers, which are estimating with our approach. Accordingly, we consider 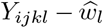, where 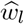 is a suitable estimator for *w*_*l*_. In regression problems with multiple dimensions such as the present case outlined in (1), the question arises as to which effects are best modelled as random versus modelled as fixed. In general, with a large number of coefficients to be estimated, the potential loss in degrees of freedom associated with modelling fixed effects is considered an argument in favor of random effects. In the large surveys considered in this article several hundred interviewers were involved in taking measurements. Estimating a fixed effect for each interviewer is thus prohibitively expensive in terms of degrees of freedom. We therefore proceed in line with common practice and assume that the household effect *u*_*j*_, and interviewer effect *w*_*l*_ are stochastic (Hodges, 2013; Hsiao, 2014; Fielding, 2004). In case of the location effect *v*_*k*_ the optimal choice is less clear. The potential loss of degrees of freedom is lower due to the lower number of coefficients to be estimated, especially at the highest level of geography. However, in order maintain maximum comparability of the level-effects, we consider it sensible to model all as random.

These random level-effects are assumed to be independently drawn from underlying normal distributions (Hodges, 2013). As part of the estimation procedure we obtain estimates for the respective second moments of these distributions, which then can be used for simulation exercises or the calculation of reliability ratios. With the assumption of random effects, equation (1) constitutes a linear mixed model (LMM), that is:

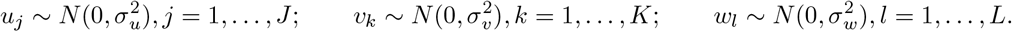

### 4.7 Omitted variable bias

An individual’s blood pressure depends on various factors, only some of which can be fully captured in large-scale surveys. Genetic preconditions for example are practically impossible to capture sufficiently in survey settings. Thus, we are agnostic about facing omitted variable bias in explaining systolic blood pressure independent of the particular survey data set considered. However, depending on the survey, some essential predictors of blood pressure are missing, which in principle could be recorded in a survey setting.

Recalling that our main interest lies in investigating interviewer effects, we are mostly concerned about falsely attributing variation in systolic blood pressure measurements to interviewers. Confounding is most likely to occur if an interviewer’s specific subset of individuals substantially differs from the overall population, along a dimension relevant for variation in systolic blood pressure.

The risk of confounded interviewer intercept estimates caused by small samples is mitigated by using the best linear unbiased predictor (BLUP) for random effects (Henderson, 1975; Rao and Molina, 2015). This estimator is a weighted average of the pooled sample and the sample from the level-specific subgroup, i.e., all measurements taken by one specific interviewer. The former exhibits a bias and small variance, whereas the latter is unbiased but has a large variance. It is constructed so that the more observations there are in the level-specific subgroup, the more weight is attributed to it. Conversely, if the level-specific subgroup sample is very small, the BLUP relies more heavily on the pooled sample. The estimation procedure therefore amounts to a variance-bias trade-off in which the BLUP is optimal in terms of the mean squared error (MSE). Consequently, the potential small sample bias that leads to confounded interviewer intercept estimates is small, and its impact negligible.

### 4.8 Testing for the Presence of Interviewer Effects

We are interested in investigating the presence and significance of interviewer effects. This relates to the formal test of the hypothesis 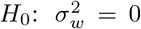 vs. 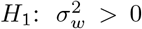. This test is performed by evaluating the likelihood-ratio statistic

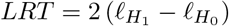

where 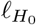 is the log-likelihood of the model under the null and 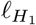 for the alternative. In our concrete case, 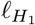 nests 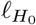 and additionally includes interviewer random effects. As fundamental problem, the null lies at the boundary of the parameter space. The asymptotic distribution of the LRT has the inconvenient distribution of a point-mass on zero with weight 0.5 and 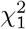-distribution elsewhere. The finite sample distributions however may severely differ from the asymptotic distribution (Crainiceanu and Ruppert, 2004, 2005). For multiple random effects as in the present model, a parametric bootstrap can approximate the finite sample distribution well enough (Crainiceanu, 2008; Greven et al., 2008). In particular,

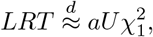

where 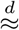 denotes approximate equality in distribution, *U* ∼ *Bern*(1 − *p*). Both *a* and *p* are unknown and have to be estimated by bootstrap replications. Eventually, p-values for the LRT under the null can be provided.

### 4.9 Adjusting for Interviewer Effects

Once we have established the presence and significance of interviewer effects, we adjust blood pressure measurements for these interviewer effects. Since we obtain not only an estimate of the second moment of the interviewer effect distribution, but also intercepts for all individual interviewers, we can individually adjust systolic blood pressure measurements. A simple adjustment then takes the form

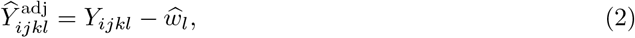

where 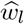 are the interviewer intercept effects (the BLUPs).

### 4.10 Assessing Uncertainty in Sample Hypertension Prevalence

In order to quantify the uncertainty in hypertension prevalence induced by interviewer measurement error we use a non-parametric bootstrap approach. Precisely, for this approach we repeatedly take sub-samples of observed systolic blood pressure measurements and their corrected counterparts and compare resulting prevalences of hypertension. We depict the two generated sets of prevalences as densities, which allows for a straightforward comparison.

#### 4.10.1 Bootstrap

We employ a non-parametric cluster bootstrap approach to infer about the uncertainty of hypertension prevalence given the corrected observations. We refer to this approach as non-parametric, since we do not use estimated parameters from the estimated model to generate new data, but only use the predicted interviewer effects to create adjusted measurements post estimation. Thus, we compare the density of hypertension prevalences based on corrected observations to the density of prevalences based on uncorrected observations. In order to account for the clustered structure of our data, we fix the coarsest geographic level (e.g. provinces) in the data and within these levels we draw from the second coarsest geographical level (e.g. municipalities).

The location level effects depict multiple levels of granularity and thus can also be represented as distinct effects. Let *p* = 1, …, *P* indicate the coarsest geographical level effect (e.g. province), and *m* = 1, …, *M* (*p*) represent the second coarsest geographical level effect (e.g. municipality).

Formally, let *y*_*ipm*_, *m* = 1, …, *M* (*p*) be the *i*th individual measurements in province *p* and 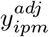 the adjusted measurements respectively. Then, *R* bootstrap replications are generated via:

~~~
1. for *r* = 1, …, *R*:
2. for *p* = 1, …, *P* :
3. Draw *M* (*p*) municipalities with replacement
4. Obtain composite sample *B*(*p*) ⊂ *{y*_*ipm*_|for individual *i* in muncipality *m}*^*M* (*p*)^
5. Pool random samples to obtain 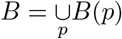
6. Calculate *p*_*r*_(*B*) = |*B*|^−1^∑_*y*∈*B*_𝕀(*y >* 140), and 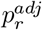 analogously
~~~

The bootstrap prevalences (*p*_*r*_)_*r*=1,…,*R*_ and 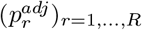 allow for inferring about the effect of adjustment.

## Data Availability

All data used in the study is available via the three health surveys they were derived from.

https://www.rand.org/well-being/social-and-behavioral-policy/data/FLS/IFLS.html

http://www.nids.uct.ac.za/nids-data/data-access

https://lasi-india.org/data-instructions

